# Incidence-weighted force of infection for predicting first reported health-zone cases during the 2026 Bundibugyo virus disease outbreak: a rolling-origin evaluation

**DOI:** 10.64898/2026.08.12.26360244

**Authors:** Johan G. L. Verheyden, Celestin Nzanzu Mudogo

## Abstract

Anticipating which health zone will report the next confirmed case is operationally distinct from forecasting national case counts and matters for prepositioning response capacity; most spatial spread models rely on mobile-phone mobility data unavailable in the Democratic Republic of the Congo (DRC).

We modelled the discrete-time hazard of a first reported confirmed case across 106 health zones in four provinces affected by the 2026 Bundibugyo virus disease outbreak (47 affected, 59 at risk, 26 July 2026), comparing four connectivity specifications — none, road-distance, a gravity score, and an incidence-weighted force-of-infection (FOI) term — fitted within an identical Bayesian hierarchical hazard architecture. Evaluation used a rolling-origin design, cluster bootstrap resampling, leave-one-origin-out and non-overlapping-origin checks, and a kernel-parameter sensitivity grid, with top-10 hit rate the pre-specified primary metric, matched to the operational question of which few zones warrant attention; AUC-PR, top-5 hit rate, and median rank percentile were secondary.

FOI had the highest top-10 hit rate (42.6%), approaching conventional significance against road-distance and no-connectivity comparators. On AUC-PR, a model with no connectivity term performed as well as or better than any connectivity specification (0.437 vs. 0.409 for FOI), a discrepancy we report rather than omit. Rankings were stable across the sensitivity grid (Spearman ρ 0.90–0.99) and across robustness checks.

An incidence-weighted connectivity term modestly and specifically improves identification of the highest-risk zones, concentrated in top-k ranking rather than uniform across metrics. The evaluation is pseudo-prospective, since historical data-vintage snapshots could not rule out retrospective revision, pending verification via a pre-registered top-20 ranking.

## 1 Introduction

The Democratic Republic of the Congo (DRC) is, as of May 2026, responding to its seventeenth recorded Ebola epidemic and only the second ever caused by Bundibugyo virus (BDBV), declared on 15 May 2026 in Ituri Province (Mwamba et al., 2026). Historically, BDBV outbreaks have been smaller and less severe than Zaire ebolavirus outbreaks in the region (Walekhwa et al., 2026; Nash et al., 2024). The 2026 outbreak has not followed that pattern: by the 26 July 2026 data cutoff used here, cumulative confirmed cases stood at 3,262 across five provinces, approaching the scale of the 2018–20 North Kivu–Ituri Zaire ebolavirus outbreak (Walekhwa et al., 2026).

Anticipating which currently unaffected health zone is likely to report the next case is a distinct, and for response prepositioning arguably more actionable, question than forecasting national case counts. A real-time dashboard combining surveillance and mobility data has been established for this outbreak (Mbulayi et al., 2026), and a separate mobility-mapping analysis found the outbreak’s epicentre zones to be among the most highly connected in the country on an independent mobility index (Bangelesa et al., 2026, as summarised in Walekhwa et al., 2026). National-level scenario projections are also available (Mooring et al., 2026). None of these efforts provides a validated, prospectively evaluated model of zone-level invasion risk specifically, and existing approaches that do address spatial spread typically rely on mobile-phone call detail record (CDR) data, which a systematic review found available in only a small number of African countries, not including the DRC (Wardle et al., 2023).

This paper develops a discrete-time hazard model in which spatial connectivity is represented by the observed incidence in every currently affected zone, propagated to at-risk zones through a road-network distance-decay kernel, rather than by geometric distance to the nearest affected zone alone. We refer to this as an incidence-weighted spatial force-of-infection (FOI) hazard model. We initially described this specification as renewal-equation-based; on reflection, that terminology overstates the model’s mechanistic content, since it lacks the temporal generation-interval convolution that a formal renewal equation requires (Cori et al., 2013), and we no longer use that terminology here. We evaluate four connectivity specifications — no connectivity term, road-distance, a gravity score, and the FOI term — fitted within an identical hazard-model architecture, using a rolling-origin design with cluster-aware uncertainty quantification, leave-one-origin-out and non-overlapping-origin checks, and a sensitivity analysis over the FOI kernel’s parameters.

Two further points of precision matter for how the results should be read. First, the outcome modelled throughout is a zone’s first reported confirmed case, not the date of biological introduction of the virus; the two can differ substantially depending on surveillance intensity, testing access, and reporting capacity, and we return to this distinction in the Discussion. Second, because historical snapshots of the surveillance dataset at each rolling-origin date were not available to us, we cannot rule out that some retrospective data revision (case reclassification, backfilled reports) is reflected in what our evaluation treats as contemporaneously known information. We therefore describe the rolling-origin evaluation as pseudo-prospective throughout, reserving the term prospective for the genuinely pre-registered future verification described in Section 2.10.

### 1.1 Relationship to companion analyses

This paper draws on the same INRB-UMIE surveillance infrastructure as two companion manuscripts from our group: a national-level short-term case-forecasting analysis (Verheyden, Nzanzu Mudogo & Jacquet, unpublished-a), evaluated against forecast-model families and interval coverage rather than spatial rank-based hit rates; and an analysis of what a health-zone case-fatality ratio measures given differential reporting capacity (Verheyden, Nzanzu Mudogo & Jacquet, unpublished-b). This paper’s outcome (zone-level hazard of first reported case), unit of analysis (health-zone-day), and evaluation framework (spatial rank-based hit rate under rolling-origin cluster bootstrap) are each distinct from both. A brief growth-rate extension for already-affected zones is summarised in Section 3.5, rather than presented as a second primary model, to keep this paper’s central contribution — the zone-level hazard comparison — clearly delineated.

## 2 Materials and Methods

### 2.1 Study design, setting, and data sources

Retrospective, pseudo-prospective observational study of health-zone-level case emergence during the 2026 BDBV outbreak, 14 May–26 July 2026 (74 days). Risk set: 106 health zones, Ituri, Nord-Kivu, Haut-Uele, Tshopo provinces. A fifth province, Sud-Kivu, reported its first case shortly before this analysis was finalised and is excluded from the risk set. Daily case data from the INRB-UMIE BDBV2026-Data repository (Mbulayi et al., 2026). Static covariates: WorldPop population estimates; OpenStreetMap/HOT health-facility points; Complex Crisis Vulnerability Index; province-level ACLED conflict events (Raleigh et al., 2010). Road-network travel times from a national OSRM graph. CDR/mobility data were not used, as DRC is not among the countries for which this is available (Wardle et al., 2023).

### 2.2 Outcome definition

The outcome modelled is a zone’s first reported confirmed case, denoted *T_z_*, with *y_z_*(*t*)=1 on the day this occurs. We use this precise language deliberately rather than ‘invasion’: first confirmation reflects viral introduction, local transmission, detection, testing access, reporting capacity, and retrospective case reclassification jointly, and the model cannot separate these. Where we use ‘invasion’ elsewhere in this paper it is shorthand for this defined outcome, not a claim about the unobserved date of biological introduction; Section 4.3 returns to the implications of this distinction, particularly for the health-facility-density covariate.

Each zone enters the risk set on day 1 and exits permanently upon its first reported case. This construction yields 5,727 zone-day observations, 47 events.

### 2.3 Statistical model: discrete-time hazard with a cloglog link

The discrete-time hazard is:

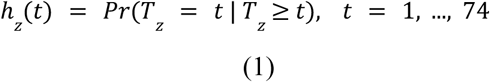

with the complementary log-log link, the discrete-time analogue of a continuous-time proportional hazards model (Prentice and Gloeckler, 1978):

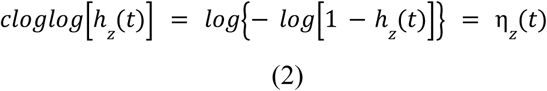

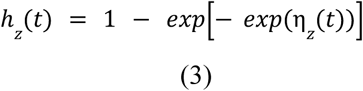

Linear predictor:

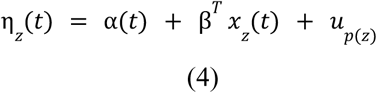

with a piecewise-constant weekly baseline evolving as a random walk:

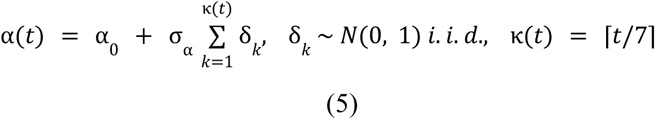

a non-centred province random effect (Betancourt and Girolami, 2015):

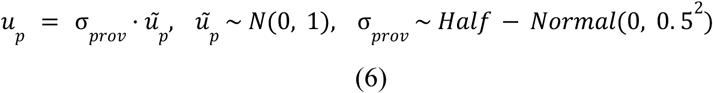

weakly informative priors:

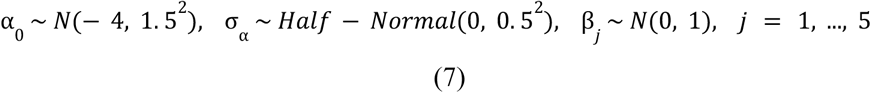

and the discrete-time survival log-likelihood, summed over zones and at-risk days:

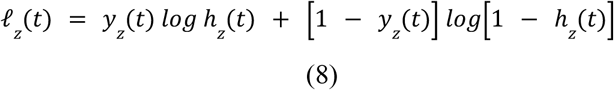

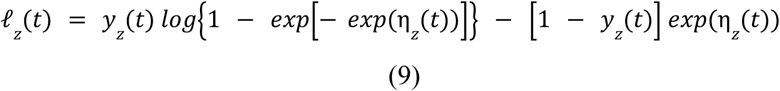

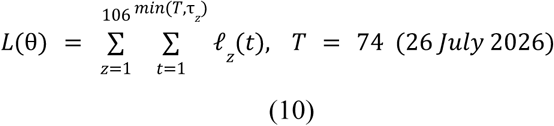

This structure is identical across all four connectivity specifications compared in this paper (Section 2.5); only the content of one covariate, *x_z_*(*t*)’s connectivity term, differs between them.

### 2.4 Data-vintage and same-day information leakage

Two distinct leakage risks apply to any covariate built from the same surveillance stream as the outcome. Same-day leakage — using zone z’’s incidence recorded on day t to predict zone z’s outcome on day t — is addressed directly: the FOI covariate (Section 2.5) uses only incidence through day t−2, never day t or t−1. Data-vintage leakage — whether the case counts available to us for a historical date t already reflect corrections made after t, since we hold only the final, most recently corrected version of the dataset rather than dated historical snapshots — cannot be ruled out with the data available to us. We flag this explicitly rather than assume it away, and for this reason describe the rolling-origin evaluation throughout as pseudo-prospective: a reconstruction of what a real-time analysis would have produced, run retrospectively on the best data now available, not a true contemporaneous test. The one exception is the prospective validation register (Section 2.9), which is a genuine pre-registration against outcomes not yet known at the time of writing.

### 2.5 Connectivity specifications, fitted within an identical architecture

Four specifications of *x_z_*(*t*)’s connectivity term were compared, all embedded in the identical model described in Section 2.3, differing only in this one covariate:

i. Baseline: no connectivity term (four static covariates only).
ii. Road-distance: log(1 + travel time to the nearest already-affected zone), the specification used in an earlier version of this analysis.
iii. Gravity: a population-weighted score summed over all affected zones:

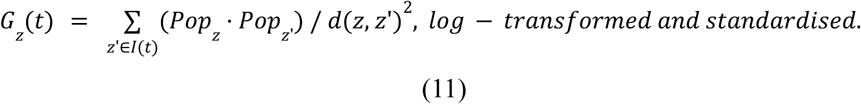

(iv) FOI (primary specification): incidence-weighted, kernel-summed force of infection,

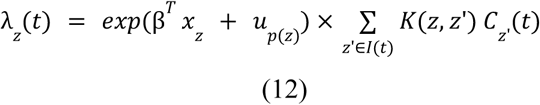

with kernel *K*(z,z’) = exp(−*d*(z,z’)/*φ*), *φ*=60 minutes (default), and *C_z’_*(*t*) the 3-day trailing mean of new confirmed cases in zone *z’* through day *t*−2 (Section 2.4). Sensitivity to *φ* and the smoothing window is reported in Section 2.9.

Fitting all four within the identical architecture, rather than comparing a fitted model against externally-computed rankings, isolates what each connectivity term contributes beyond the static covariates and beyond each other, and was added specifically in response to reviewer concern that an earlier version compared a fitted model to uncalibrated external scores.

### 2.6 Bayesian estimation

PyMC 6.2.0 (Abril-Pla et al., 2023), No-U-Turn Sampler (Hoffman and Gelman, 2014), 4 chains × 1,000 post-warm-up draws (1,500 tuning), target acceptance 0.92–0.95, seed 20260729. All four variants converged (max R-hat 1.00–1.01, 1–3 divergent transitions of 4,000 draws each). Static covariates standardised prior to fitting.

### 2.7 Rolling-origin evaluation: primary and secondary metrics

Eight weekly origins, 21 May–9 July 2026, each refit using only data available through that origin, scored against zones reporting a first case within the following 14 days (Tashman, 2000). We designate top-10 hit rate — the proportion of true next-invading zones ranked in the model’s top 10 at that origin — as the primary evaluation metric, specified before examining comparative results across specifications, because the operational decision this model is built to support is which small number of zones warrant prioritised attention, not discrimination across the full 59-zone risk set. AUC-PR (Saito and Rehmsmeier, 2015), top-5 hit rate, and median rank percentile of the true next-invading zone are reported as secondary metrics. Where these diverge from the primary metric’s conclusion — which occurs for AUC-PR (Section 3.2) — we report the divergence rather than the primary metric alone.

### 2.8 Cluster bootstrap, leave-one-origin-out, and non-overlapping-origin checks

Zone-level scores within one origin share a single model fit and are not independent; a naive bootstrap over zone-rows would understate uncertainty. We used a cluster (block) bootstrap resampling whole origins with replacement (10,000 resamples), for each replicate pooling the zone-level rows (or origin-level AUC-PR values) belonging to the resampled origin set and recomputing each metric as in the point estimate. We report 95% percentile intervals on each method’s metrics and on paired differences between methods, computed from the same resampled origin set.

Because the 14-day evaluation horizons overlap between origins spaced 7 days apart, adjacent origins share training data, forecast periods, and some invasion events; the cluster bootstrap addresses correlation between zone-scores within an origin but not fully between adjacent origins. We report two further checks: leave-one-origin-out (recomputing pooled metrics excluding each origin in turn, to assess whether any single origin drives the result), and a non-overlapping-origin subset (4 origins spaced exactly 14 days apart: 21 May, 4 June, 18 June, 2 July), which removes horizon overlap entirely at the cost of halving the usable origins and events (61 to 35).

### 2.9 Sensitivity analysis: FOI kernel parameters

The FOI kernel’s decay scale (φ, fixed at 60 minutes in the primary specification) and smoothing window (3 days) were not tuned against performance; we instead report ranking stability across a grid of plausible alternatives, refitting the full model at each combination and comparing the resulting zone ranking to the primary specification via Spearman rank correlation and top-10 overlap: φ ∈ {30, 60, 120} minutes × window ∈ {1, 3, 7} days (8 combinations in addition to the primary specification).

### 2.10 Prospective validation register

The FOI model’s top-20 zone ranking (26 July 2026 cutoff), SHA-256 checksums of the ranking file, and a pre-specified verification protocol — checking at 9 and 16 August 2026 whether any top-ranked zone reports a first case, cross-checked against situation reports — were fixed in a dated register before the outcome was known. This is the genuinely prospective element of this paper; results were not available at the time of writing.

### 2.11 Growth-rate extension for already-affected zones (summary)

A secondary, exploratory analysis — not a second validated model of comparable standing to the hazard model above — examined near-term case growth for the 47 zones already affected, using a hierarchical negative-binomial growth-rate model (an initial Poisson specification was found miscalibrated on rolling-origin backtesting and corrected). We summarise the headline finding here (Section 3.5) and do not present a combined invasion-plus-growth ranking as a validated decision tool in the main paper, since that combined ranking has not itself been backtested.

### 2.12 Software and reproducibility

Python 3.12, PyMC 6.2.0 (Abril-Pla et al., 2023), ArviZ, NumPyro (rolling-origin refits), GeoPandas, scikit-learn (AUC-PR). Code, derived data, bootstrap replicates, and backtesting results are available from the corresponding author upon reasonable request and will be deposited, with a versioned release and commit hash, upon acceptance.

## 3 Results

### 3.1 Descriptive overview

As of 26 July 2026, 47 of 106 risk-set zones (44.3%) had reported at least one confirmed case; 59 remained at risk. Fig 1 maps the risk set under the FOI model’s ranking.

**Fig 1.**
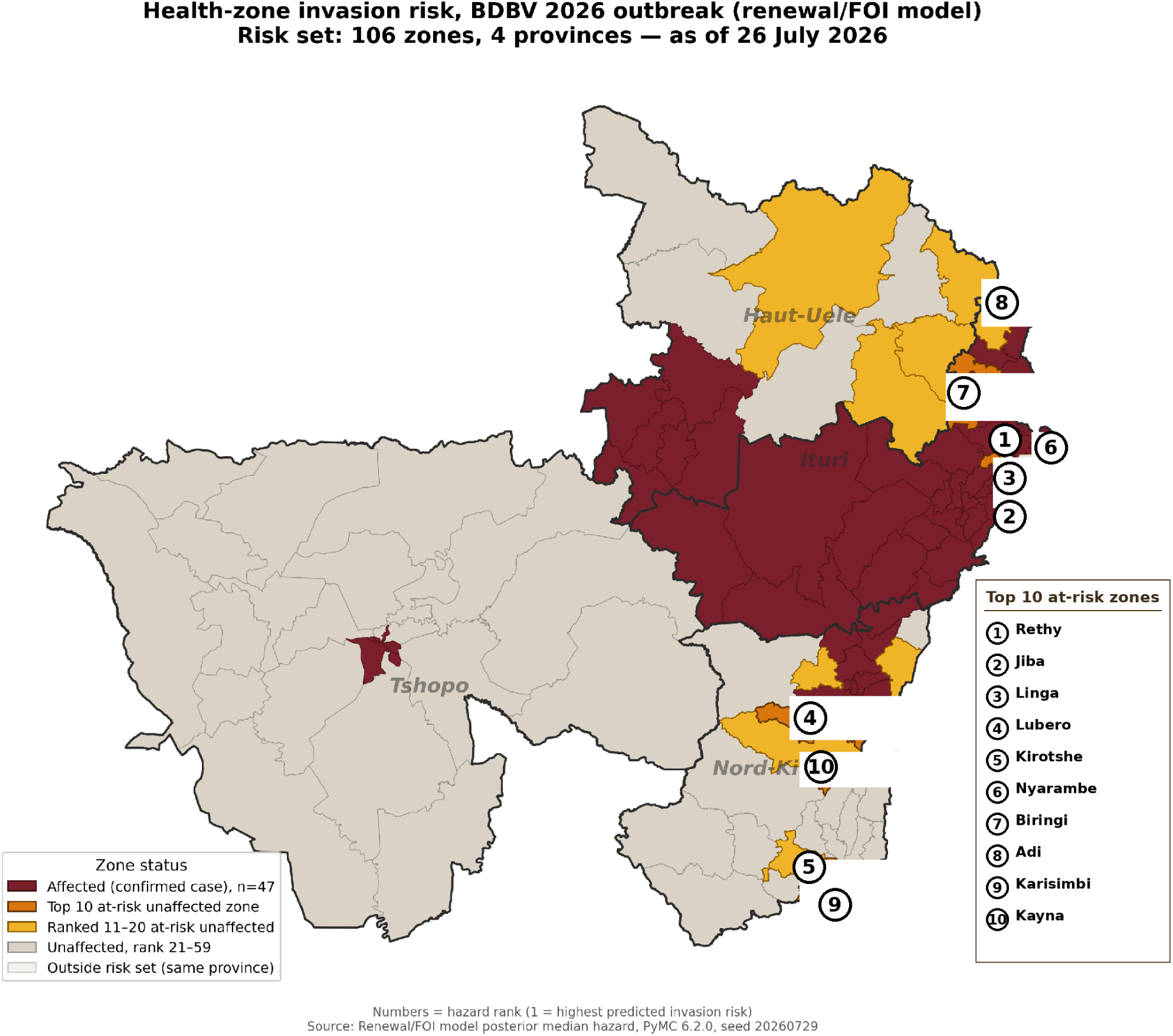
Health-zone hazard map, incidence-weighted FOI model. Affected zones (n=47) in maroon; at-risk zones shaded by hazard rank, top 10 individually labelled.

### 3.2 Architecture-matched comparison: primary and secondary metrics

Table 1 and Fig 2 report all four connectivity specifications, fitted within the identical hazard architecture, evaluated by rolling-origin design with cluster-bootstrap 95% CIs. On the pre-specified primary metric (top-10 hit rate), FOI was highest (42.6%), followed by OSRM-distance and gravity (both 32.8%), then baseline (29.5%). The paired comparison against OSRM-distance reached the boundary of conventional significance (+0.105, 95% CI [0.000, 0.244]); against gravity and against baseline it was directionally favourable but did not clear zero (gravity: +0.101, CI [−0.032, 0.256]; baseline: +0.132, CI [−0.019, 0.302]).

**Fig 2.**
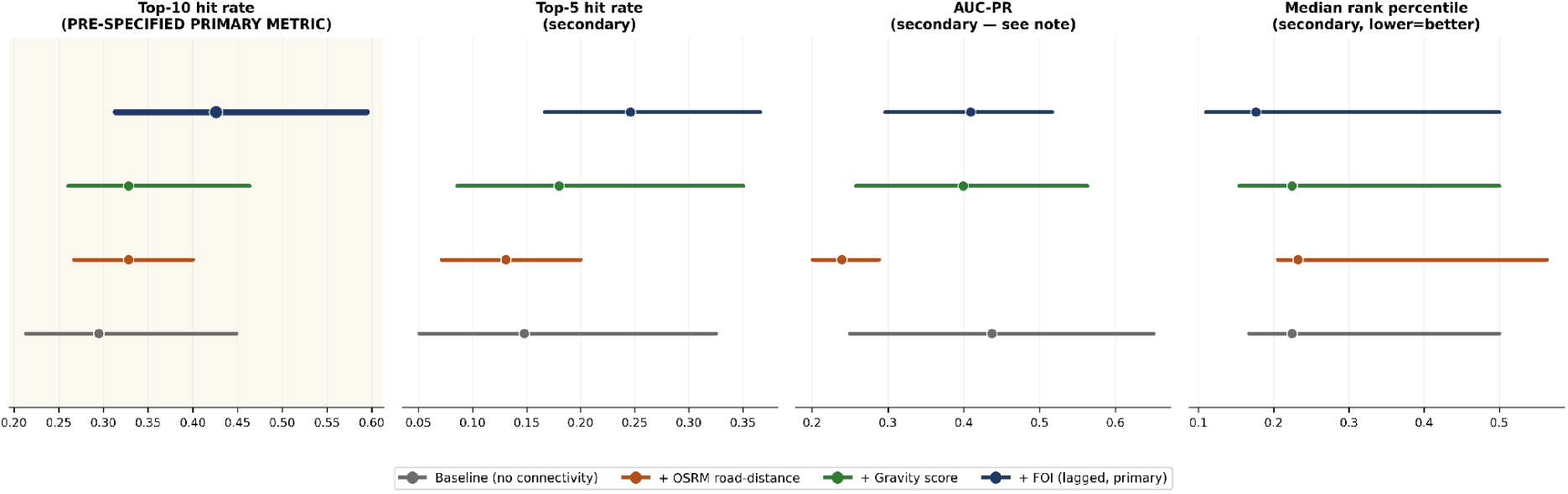
Architecture-matched rolling-origin comparison, cluster-bootstrap 95% CIs. All four variants fit within the identical hazard model; only the connectivity covariate differs. Top-10 hit rate (shaded panel) is the pre-specified primary metric.

**Table 1.**
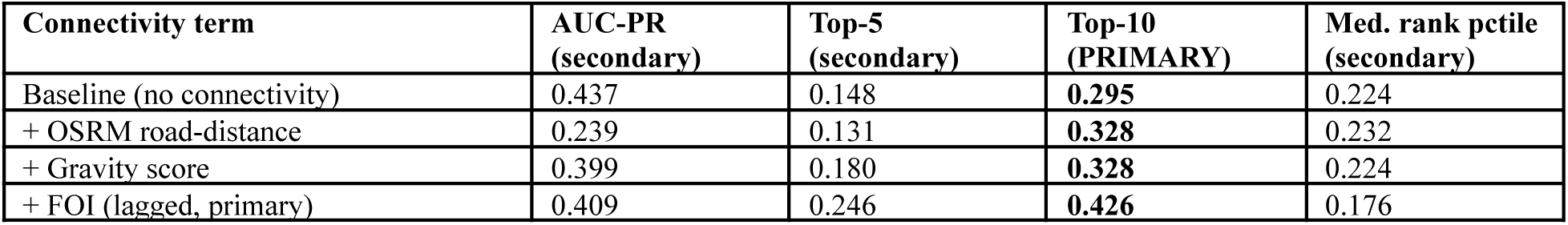
Architecture-matched comparison, all four connectivity specifications, rolling-origin evaluation (point estimates; see Fig 2 for bootstrap 95% CIs).

On AUC-PR, the secondary metric with the most divergent picture, the baseline model with no connectivity term at all performed as well as or better than every connectivity specification (baseline 0.437 vs. FOI 0.409, gravity 0.399, OSRM-distance 0.239), and FOI’s advantage over baseline on this metric was not distinguishable from zero (−0.027, 95% CI [−0.274, 0.200]). We report this plainly rather than selectively: a global discrimination metric like AUC-PR rewards a model for correctly ranking the bulk of zones that will not invade soon, which the static covariates alone (population, facility density, deprivation) appear to do reasonably well without any connectivity information; the connectivity terms’ distinguishable contribution is specifically to sharpening the top of the ranking, which is what the primary metric measures and what the operational use case (which few zones get attention first) requires. Table 1 gives the complete picture across both primary and secondary metrics so readers can weigh this differently if their use case differs from ours.

### 3.3 Robustness: leave-one-origin-out, non-overlapping origins, kernel sensitivity

Leave-one-origin-out: FOI’s top-10 hit rate ranged 39.3–48.0% and AUC-PR ranged 0.377–0.444 across the 8 folds excluding one origin at a time — no single origin drove the headline result. The non-overlapping 4-origin subset (61 invasion events reduced to 35, but with 14-day horizons no longer sharing data between origins) reproduced the full 8-origin FOI numbers closely (top-10 hit rate 42.6% vs. 42.9%; AUC-PR 0.409 vs. 0.415), evidence the result is not an artefact of origin overlap.

The FOI kernel’s decay scale and smoothing window were not tuned against performance. Across a grid of 8 alternative combinations (φ ∈ {30,60,120} minutes × window ∈ {1,3,7} days), the resulting zone ranking correlated with the primary specification’s ranking at Spearman ρ = 0.90–0.99, with top-10 overlap of 9 or 10 out of 10 zones in every combination tested.

### 3.4 Covariate effects, FOI specification

Table 2 (Section 3.6) and Fig 3 summarise posterior covariate effects. FOI itself: β = +0.357, 95% CrI [0.16, 0.55]. This is smaller than an earlier, same-day (non-lagged) specification’s estimate of +0.610 [0.43, 0.79]; fixing same-day information leakage (Section 2.4) reduced the coefficient’s magnitude by roughly 40%, confirming the leakage had inflated the apparent effect, though the corrected estimate still excludes zero. Health-facility density (β = +0.486, [0.245, 0.734]) and population (β = +0.400, [0.087, 0.720]) were also associated with hazard at the 95% credible level; socioeconomic deprivation (+0.326, [−0.074, 0.754]) and province-level conflict exposure (+0.156, [−0.640, 0.900]) were not.

**Fig 3.**
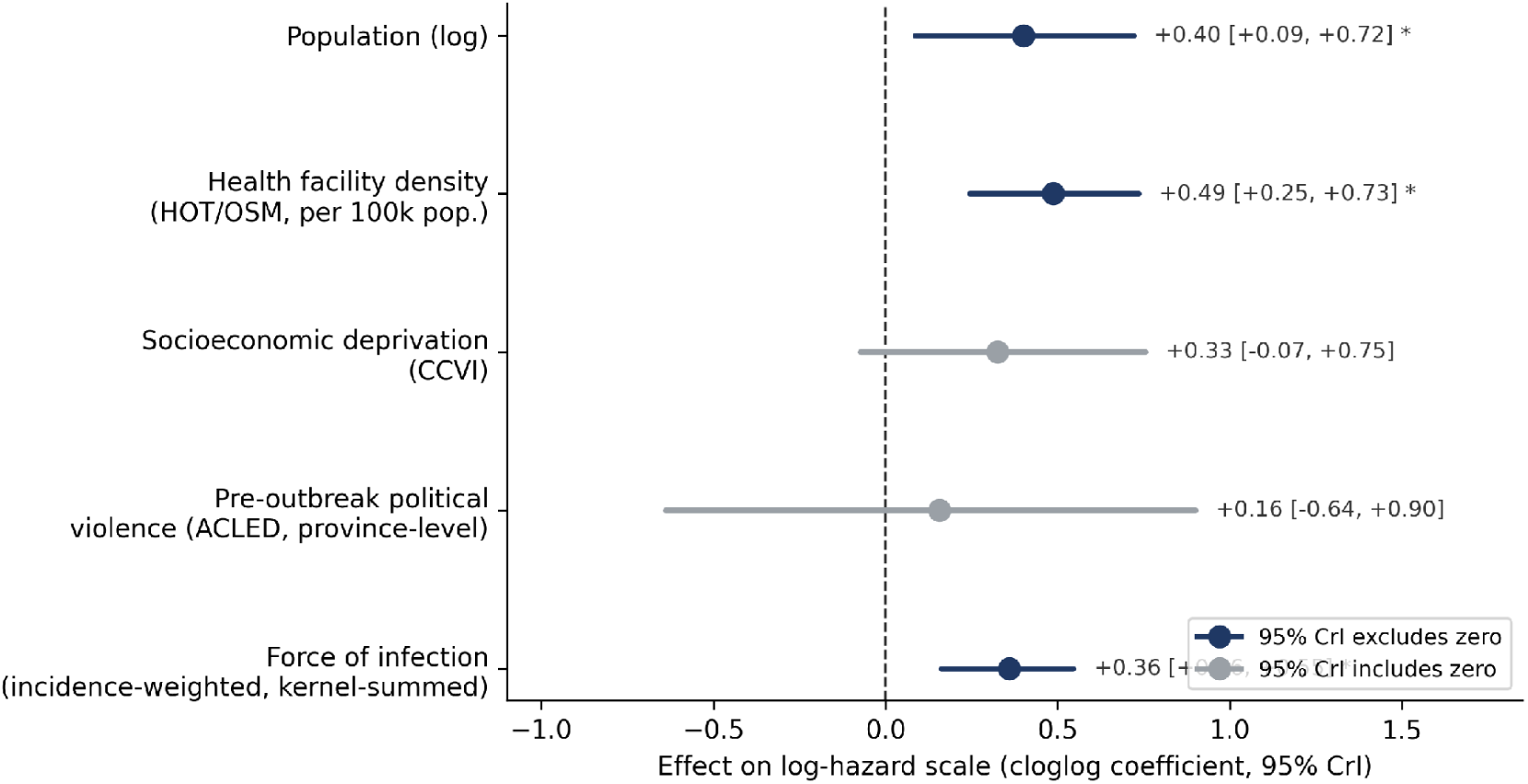
Covariate effects on hazard of first reported case, FOI model. Posterior medians and 95% credible intervals.

**Table 2.** Top 20 at-risk health zones ranked by posterior median hazard of first reported case, FOI model, 26 July 2026.

| Rank | Zone | Province | Daily hazard (median) | 95% CrI | Chance of 1st case in 14d |
| --- | --- | --- | --- | --- | --- |
| 1 | Rethy | Ituri | 0.0948 | [0.0181, 0.2947] | 75% |
| 2 | Jiba | Ituri | 0.0385 | [0.0081, 0.1180] | 42% |
| 3 | Linga | Ituri | 0.0337 | [0.0077, 0.0934] | 38% |
| 4 | Lubero | Nord-Kivu | 0.0250 | [0.0054, 0.0760] | 30% |
| 5 | Kirotshe | Nord-Kivu | 0.0174 | [0.0035, 0.0540] | 22% |
| 6 | Nyarambe | Ituri | 0.0158 | [0.0032, 0.0507] | 20% |
| 7 | Biringi | Ituri | 0.0134 | [0.0030, 0.0353] | 17% |
| 8 | Adi | Ituri | 0.0128 | [0.0026, 0.0399] | 17% |
| 9 | Karisimbi | Nord-Kivu | 0.0124 | [0.0019, 0.0509] | 16% |
| 10 | Kayna | Nord-Kivu | 0.0114 | [0.0026, 0.0331] | 15% |
| 11 | Mutwanga | Nord-Kivu | 0.0092 | [0.0021, 0.0250] | 12% |
| 12 | Watsa | Haut-Uele | 0.0090 | [0.0017, 0.0321] | 12% |
| 13 | Laybo | Ituri | 0.0087 | [0.0018, 0.0245] | 12% |
| 14 | Angumu | Ituri | 0.0067 | [0.0013, 0.0223] | 9% |
| 15 | Alimbongo | Nord-Kivu | 0.0052 | [0.0011, 0.0144] | 7% |
| 16 | Dungu | Haut-Uele | 0.0041 | [0.0008, 0.0133] | 6% |
| 17 | Masisi | Nord-Kivu | 0.0039 | [0.0008, 0.0114] | 5% |
| 18 | Biena | Nord-Kivu | 0.0035 | [0.0007, 0.0115] | 5% |
| 19 | Aba | Haut-Uele | 0.0035 | [0.0007, 0.0112] | 5% |
| 20 | Makoro | Haut-Uele | 0.0035 | [0.0006, 0.0113] | 5% |

### 3.5 Growth-rate extension (summary)

The exploratory growth-rate analysis for already-affected zones (Section 2.11) found, after correcting an initial Poisson likelihood misspecification with a negative-binomial specification, that pooled 95% interval coverage on rolling-origin backtesting rose from 33.3% (7-day horizon) and 52.1% (14-day horizon) under the original specification to 98.8% and 97.6% respectively under the corrected one (165 zone-origin backtest observations, 5 historical origins; Fig 4). Two backtest observations (Goma, Aungba) reflect retrospective downward case-count corrections in the source data rather than negative disease incidence; both are retained rather than excluded and are marked explicitly in Fig 4. We do not present a combined invasion-and-growth ranking, since that combined ranking has not itself been backtested.

**Fig 4.**
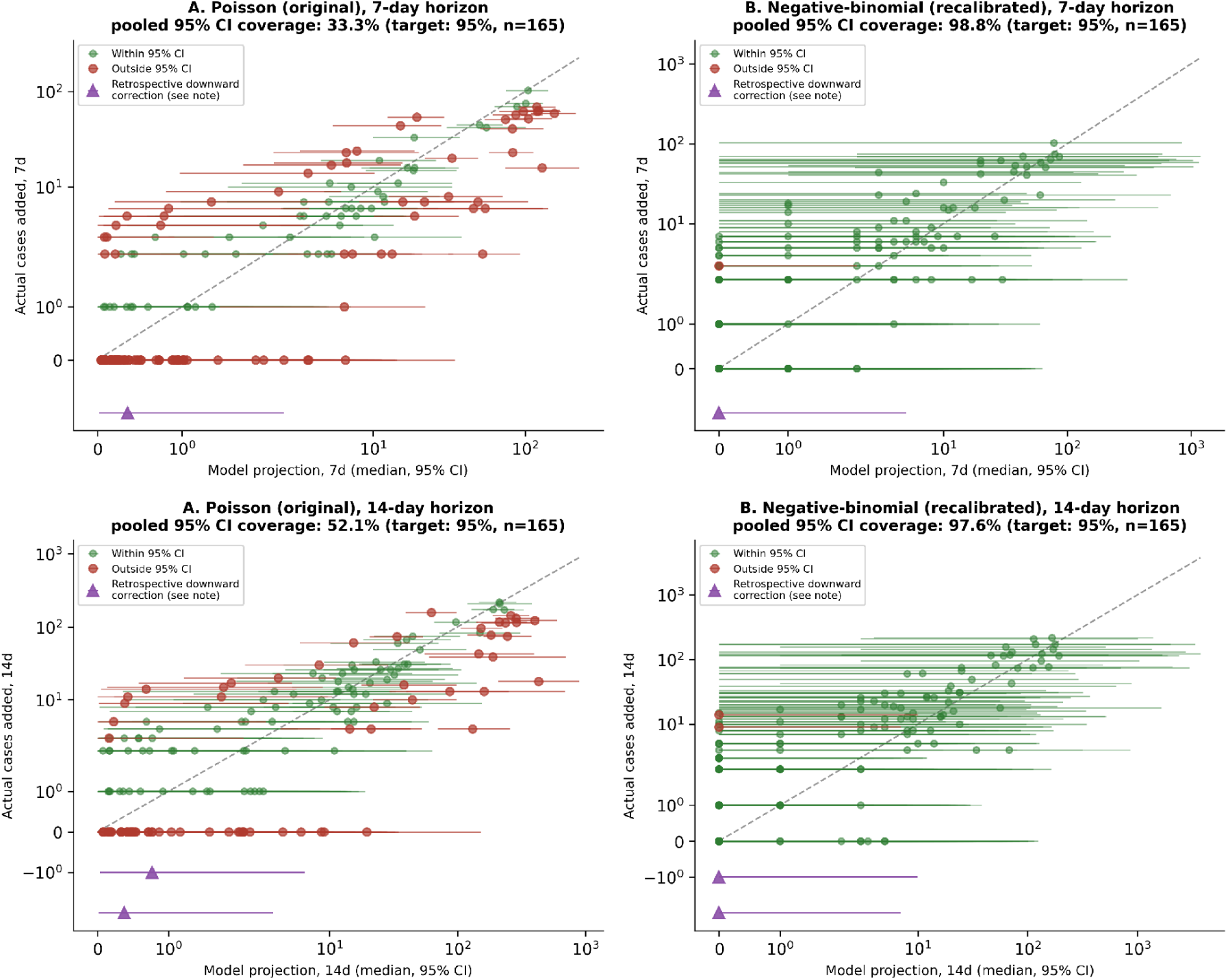
Rolling-origin backtest calibration, growth-rate extension, both horizons. Poisson (original) vs. negative-binomial (corrected) likelihood. Purple markers: retrospective data corrections.

### 3.6 Highest-risk zones

Table 2 lists the twenty at-risk zones ranked highest by the FOI model’s posterior median hazard. The top-ranked zone, Rethy (Ituri), had a posterior median daily hazard of 0.095 (95% CrI [0.018, 0.295]), corresponding to roughly a 75% chance of reporting a first case within 14 days if current conditions persisted; the 20th-ranked zone’s daily hazard corresponds to roughly 4% over the same window. Fig 5 shows daily hazard trajectories for the current top-10 zones across the full observation window.

**Fig 5.**
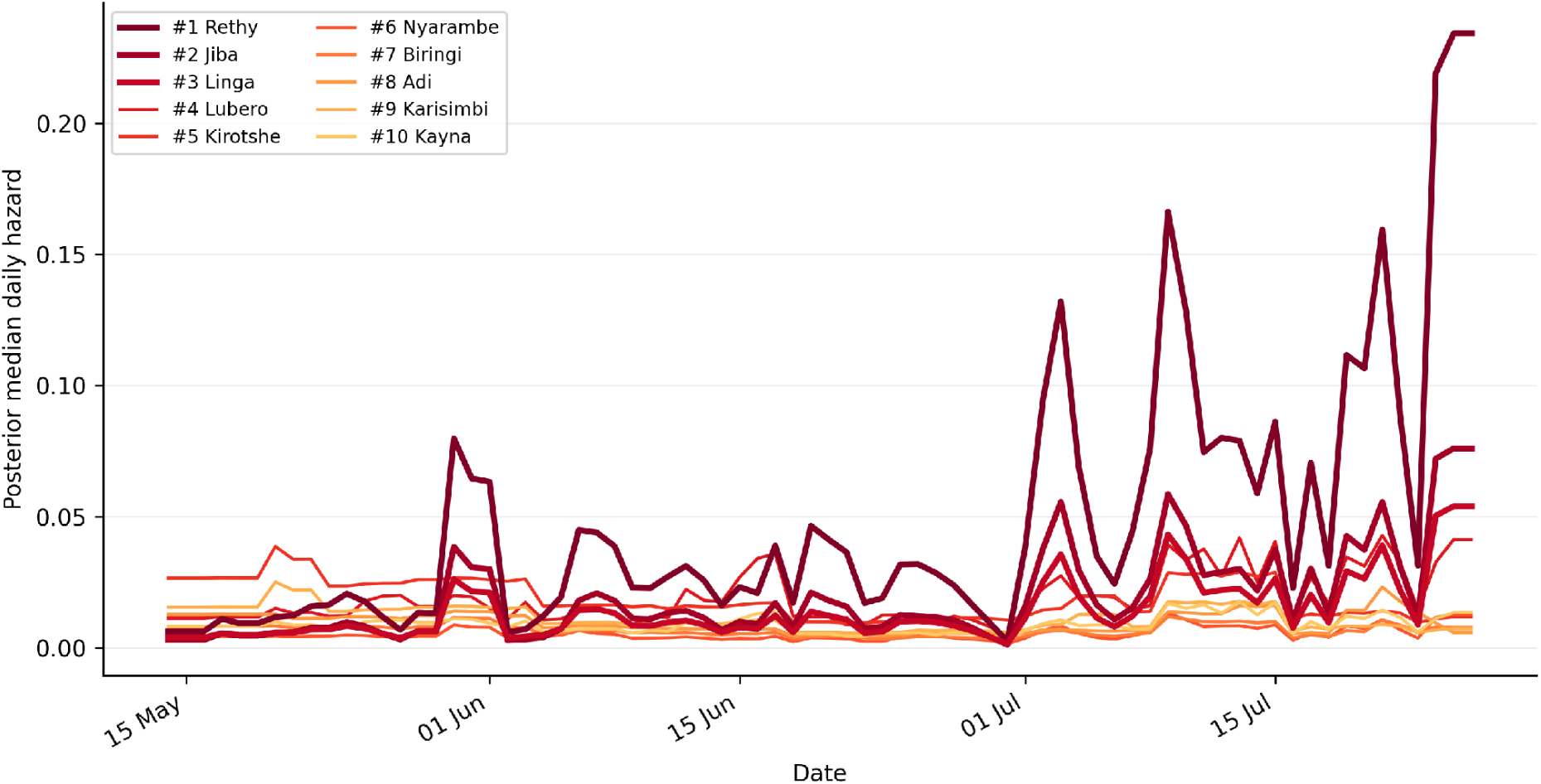
Daily hazard trajectories, top-10 at-risk zones, FOI model, 14 May–26 July 2026.

## 4 Discussion

### 4.1 Principal findings

An incidence-weighted spatial force-of-infection term, fitted within an identical hazard-model architecture, produced the highest top-10 hit rate of four connectivity specifications compared, with the improvement over a naive road-distance benchmark approaching conventional significance under cluster-aware uncertainty quantification. This advantage was specific to the top of the ranking; on a global discrimination metric (AUC-PR), a model with no connectivity term at all performed comparably or better. We regard the top-10 hit rate result as the more operationally relevant finding, since it is the pre-specified primary metric matched to the actual decision this model is meant to support, but we report the AUC-PR divergence in full because it materially qualifies how strong a claim the data support.

### 4.2 Why connectivity terms diverge on primary vs. secondary metrics

A plausible mechanism: the static covariates (population, facility density, deprivation) appear to carry broad, geography-correlated signal about which zones are generically more or less likely to be reached by a spreading outbreak, sufficient to rank the bulk of low-risk zones correctly without any connectivity information, which AUC-PR rewards across the full ranking. The connectivity terms’ distinguishable value is concentrated at the top of the ranking — correctly identifying the handful of zones at genuinely elevated near-term risk — which the static covariates alone do less well, and which top-k hit-rate metrics are specifically sensitive to. We did not anticipate this divergence and think it is a more informative finding than either metric reported alone would have been.

### 4.3 Detection versus biological introduction

The outcome modelled throughout is first reported confirmed case, not the unobserved date of viral introduction. These differ to the extent that introduction, local transmission, detection, testing access, and reporting capacity differ across zones, and we cannot separate these mechanisms with the data available. This matters specifically for the health-facility-density covariate, whose positive association with hazard may partly or wholly reflect greater detection capacity in better-served zones rather than, or in addition to, genuinely greater introduction risk. We are not aware of a way to fully resolve this with routinely available surveillance data; a first-suspected-case or first-notification outcome, where reliably available, would be a natural sensitivity analysis for future work.

### 4.4 Triangulation with independent mobility-based evidence

An independent Flowminder-based mobility analysis found the outbreak’s three epicentre zones among the top 5% of DRC health zones nationally on mobility intensity (Bangelesa et al., 2026, as summarised in Walekhwa et al., 2026), using data and methodology with no overlap with ours. That a directly measured mobility signal and our structural, incidence-weighted road-network signal both implicate connectivity in the outbreak’s early spread is external, if indirect, support for the general premise, independent of this paper’s own comparative evaluation.

### 4.5 Outbreak severity

The current outbreak’s raw case fatality ratio (44.0% at the 26 July cutoff) exceeds the historical BDBV range reported in prior meta-analysis (Nash et al., 2024); a companion analysis examines case-fatality measurement and ascertainment heterogeneity for this outbreak in detail (Verheyden et al., unpublished-b), and we do not repeat that analysis here.

### 4.6 Limitations

Data-vintage leakage (Section 2.4) cannot be excluded; the evaluation is pseudo-prospective, and the genuinely prospective element is limited to the registered top-20 ranking (Section 2.10), whose check-in results were not available at the time of writing. The top-10-vs-OSRM-distance comparison reaches only the boundary of conventional significance (CI lower bound 0.000); we would not describe this as a robust result on its own and rely on the convergent evidence across the primary metric, the sensitivity grid, and the leave-one-origin-out and non-overlapping-origin checks collectively, rather than on any single comparison. Conflict exposure is measured only at province level across four provinces, alongside a province random effect estimated from the same four groups; this covariate’s estimate should be read as exploratory. The growth-rate extension (Section 3.5) has not been backtested as a combined ranking with the hazard model, and we do not present one for that reason. Fixed choices not evaluated here include the OSRM travel-time transformation and the province-level (rather than zone-level) conflict aggregation.

### 4.7 Strengths and future work

The architecture-matched comparison, cluster bootstrap, leave-one-origin-out, non-overlapping-origin, and kernel sensitivity checks were each added specifically to close a route by which this paper’s central comparison could otherwise be an artefact of comparator choice, origin dependence, or arbitrary fixed parameters; the ranking held up across all of them. Future work should incorporate the validation register’s outcomes once available, and, if historical data-vintage snapshots become accessible, re-run the pseudo-prospective evaluation as genuinely prospective.

## Conclusions

An incidence-weighted spatial force-of-infection specification, evaluated against three alternatives within an identical hazard-model architecture, modestly and specifically improves identification of which few health zones are at highest near-term risk of a first reported case, though this advantage is concentrated in top-k ranking performance rather than uniform across every evaluation metric, and should be read as pseudo-prospective evidence pending genuinely prospective verification via the registered check-ins.

## Author Contributions

Conceptualization: JGLV. Data curation: JGLV. Formal analysis: JGLV. Methodology: JGLV. Software: JGLV. Writing – original draft: JGLV. Writing – review & editing: JGLV, CNM.

## Ethics Statement

This analysis uses publicly available, aggregate, de-identified surveillance data. [Placeholder — author input required: insert formal ethics exemption statement from the appropriate institutional review body, or a statement that this study is exempt from full ethics review, per journal policy.]

## Data Availability

Derived data and code: available from the corresponding author upon reasonable request, with public deposit, versioned release, and commit hash to follow upon acceptance. Underlying surveillance data: INRB-UMIE repository (Mbulayi et al., 2026). [Placeholder — author input required: cite the INRB-UMIE repository’s terms of use explicitly, including licence type, and list WorldPop, OpenStreetMap/HOT, and administrative boundary licences with version/access dates.]

## Acknowledgments

We thank the field research network of Aries Consult for logistical and contextual support, and the INRB-UMIE data coordination team.

## Declaration of generative AI and AI-assisted technologies in the manuscript preparation process

During the preparation of this work the author(s) used Claude (Anthropic) in order to assist with manuscript drafting, formatting, and reference organisation. After using this tool/service, the author(s) reviewed and edited the content as needed and take full responsibility for the content of the published article.

